# Risk of severe outcomes among Omicron sub-lineages BA.4.6, BA.2.75 and BQ.1 compared to BA.5 in England

**DOI:** 10.1101/2023.07.14.23292656

**Authors:** Giulia Seghezzo, Sophie G Nash, Nurin Abdul Aziz, Russel Hope, Jamie Lopez Bernal, Eileen Gallagher, Gavin Dabrera, Simon Thelwall

## Abstract

Since the emergence of Omicron variant of SARS-CoV-2 in late 2021, a number of sub-lineages have arisen and circulated internationally. Little is known about the relative severity of Omicron sub-lineages BA.2.75, BA.4.6 and BQ.1. We undertook a case-control analysis to determine the clinical severity of these lineages relative to BA.5, using whole genome sequenced, PCR-confirmed infections, between 1 August 2022 to 27 November 2022, among those who presented to emergency care in England 14 days after and up to one day prior to the positive specimen. A total of 10,375 episodes were included in the analysis, of which 5,207 (50.2%) were admitted to hospital or died. Multivariable conditional regression analyses found no evidence for greater odds of hospital admission or death among those with BA.2.75 (OR= 0.96, 95% CI: 0.84 to 1.09), and BA.4.6 (OR= 1.02, 95% CI: 0.88 to 1.17) or BQ.1 (OR= 1.03, 95 % CI: 0.94 to 1.13) compared to BA.5. Future lineages may not follow the same trend and there remains a need for continued surveillance of COVID-19 variants and their clinical outcomes to inform the public health response.

## Introduction

The Omicron variant of SARS-CoV-2 (B.1.1.529) was first detected in November 2021 in Botswana, and soon gained global dominance over the previously dominant Delta variant. (1,2) Since then, several sub-lineages of Omicron have arisen and continuously displaced the previous sub-lineage in the UK. (3) In April 2022, the Omicron sub-lineage BA.5 emerged and by June 2022 had taken dominance over BA.1 and BA.2 in the UK. BA.5 has spike protein mutations similar to that of BA.1 and BA.2, in addition to others such as, L452R and F486V; raising concerns that this sub-lineage had the potential for immune evasion to vaccines and therapeutic agents. (4)

Several other sub-lineages have since emerged, with BA.2.75 being declared as a variant of concern in England on July 18, 2022, having first been detected in the country in June 2022. It has a reversion in the gene encoding the spike protein: R493Q, similar to that seen in BA.5.(5) After BA.2.75, the sub-lineage BA.4.6 was declared a variant of concern on September 1, 2022, in England, having first been detected in the country on May 12, 2022. The BA.4.6 sub-lineage acquired a mutation in spike: R346T, a site that has potential antigenic significance. (5) The most recent of the three sub-lineages to be declared a variant of concern was BQ.1, declared in October 2022 after being first detected in England in April 2022. This sub-lineage has acquired spike mutations L452R, and K44T, along with R346T as seen with previous sub-lineages, which is associated with a growth advantage.(5,6) All three Omicron sub-lineages, BA.4.6, BA.2.75 and BQ.1, carry the N460K mutation. *In vitro* studies suggest that this mutation may confer increased immune evasion, which could lead to increased transmissibility and potentially increased in disease severity. (6,7) BQ.1 has a reported growth advantage of 38.6% (95% Confidence Interval (CI): 33.9 to 44.0) relative to BA.5.2, this significant growth advantage has also been observed in other countries and quickly became the most dominant variant in some regions globally.(3)

Previous separate studies have indicated that the infection severity for Omicron is lower than for Delta, with BA.2 showing slightly lower severity than BA.1 and the BA.4 and BA.5 sub-lineages showing similar severity to BA.2. (8) There is currently limited information about the severity of disease following infection with the most recent lineages of BA.4.6, BA.2.75 and BQ.1, with some evidence suggesting there is no difference in severity between BQ.1 and BA.5. (9,10)

## Methods

### Study design

We undertook a case-control study which included all episodes of PCR-confirmed SARS-CoV-2 infection in England, between 1 August 2022 and 27 November 2022 for which Whole Genome Sequencing indicated infection with Omicron sub-lineages BA.4.6, BA.5, BA.2.75 or BQ.1.1; these were restricted to those which occurred among individuals who attended a National Health Service (NHS) emergency department (ED) between one day before their positive test date and 14 days after their positive test date. A second population was used for sensitivity purposes, including only individuals who tested positive on the day of ED attendance.

Individuals were excluded if covariate information on age, sex, and vaccination status was missing. Healthcare workers identified via the SARS-CoV-2 immunity and reinfection evaluation (SIREN) study, which involves regular screening of healthcare workers whose test results thus are not representative of infection in the wider population eligible for PCR testing, were also excluded. (11)

### Data sources

Individual-level data on laboratory-confirmed COVID-19 infections were extracted from the Second Generation Surveillance System (SGSS), the national surveillance system that holds test results from diagnostic laboratories in England for notifiable infectious disease, and linked to validated WGS data coordinated by the COVID-19 Genomics UK consortium, available on the Cloud Infrastructure for Big Data microbial Bioinformatics database, using a unique identifier. (12,13)

UKHSA defines variants as genomes that contain a set of defining mutations, allowing consistent detection, monitoring and surveillance of cases. UKHSA variants are not equivalent to lineages assigned by Pangolin. (13) The sequence data used in this analysis is classified using the UKHSA variant definitions for V-22APR-04 (BA.5), V-22SEP-01 (BA.4.6), V-22JUL-01 (BA.2.75) and V-22OCT-01 (BQ.1.1). Further information about variant definitions used in this analysis can be found in the supplementary materials.

COVID-19 data were then linked to the Emergency Care Data Set (ECDS) and the Secondary Uses Service (SUS) dataset to obtain data on hospital attendance and admission. (15) Vaccination status was determined using data from the National Immunisation Management Service (NIMS). (16) Data on COVID-19 deaths was sourced from the UKHSA COVID-19 mortality dataset. (17)

Age, sex and area of residence were extracted from SGSS. National-level Index of Multiple Deprivation (IMD), an area-level measure of relative socioeconomic deprivation, were assigned using the lower super output area (LSOA) of residence association with the first positive specimen within an episode, using a 2019 LSOA lookup from the Office for National Statistics. Ethnicity was determined using either self-reported ethnicity through Pillar 2 testing or via linkage to the NHS Digital Hospital Episode Statistics Admitted patient care, Accident and Emergency and Outpatient databases, using categories as classified by the 2001 ONS census. (12)

#### Outcome and adjustment variable definitions

The outcome used in this analysis has been previously used to assess severity and it is defined as individuals having attended the ED and being transferred or admitted to hospital and having a length of stay in hospital for 2 or more days; or those who died during their ED attendance or up to 2 days after their initial date of ED attendance.(18) These cases were compared with a control group of COVID-19 patients whose ED attendance ended with discharge, and who did not die in the 2 days following ED attendance.

Two population definitions were used in the analysis. Population 1 were individuals who attended a NHS ED between one day before positive test date and 14 days after their positive test date, as previously described above. Population 2 included only individuals who tested positive on the day of ED attendance. The main analyses were performed using population 1, and population 2 was used for sensitivity purposes.

Vaccination status was defined as the time since last vaccination received at least 14 days before an individual’s positive test. This included any vaccine dose including boosters.

#### Statistical analysis

Analyses were run separately for confirmed cases of BA.4.6, BA.2.75 or BQ.1.1 against BA.5. Odds ratios (OR) and 95% confidence intervals (CI) were estimated using conditional logistic regression. Each of the three models were stratified for week of positive test, and further adjusted for age using categorical 10-year age bands, sex, vaccination status and NHS England region.

## Results

Between 1 August 2022 and 27 November 2022 a total of 10,820 people had a sequenced PCR positive COVID-19 sample and attended the ED according to the criteria described previously. 445 episodes were excluded from the study (433 missing covariate information and 2 from SIREN study). Throughout the study period, BA.4.6 episodes were steady between August and October 2022, while BQ.1. and BA.2.75 episodes rose during October and November 2022 (Figure 1).

**Figure 1:**
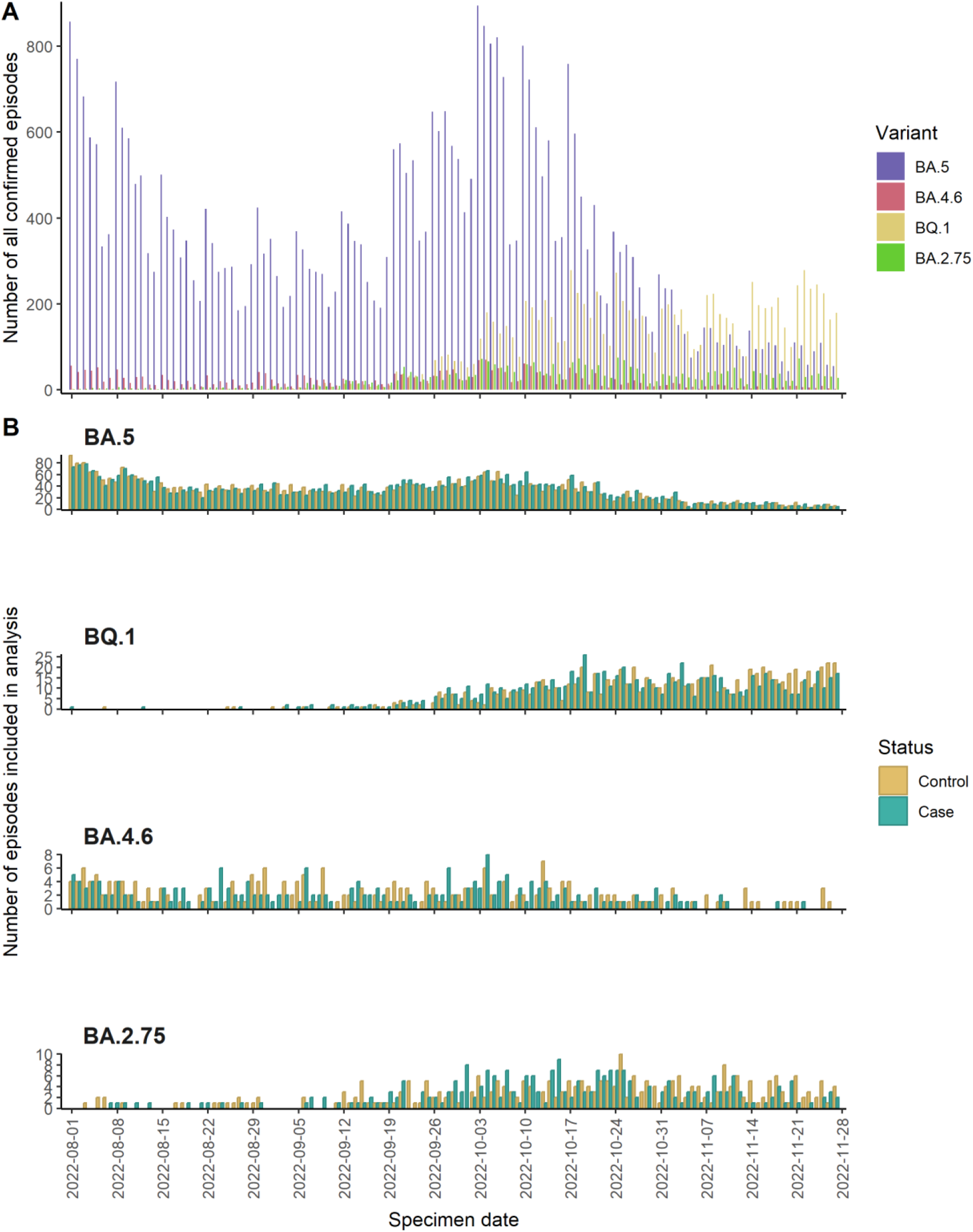
Epicurves of cases and controls for BA.5, BA.4.6, BA.2.75 and BQ.1

A total of 10,375 episodes met the study criteria for the primary population, Population 1, and were included in the analysis, of which 5,207 (50.2%) were admitted or died. A total of 1,564 cases and controls were included for BQ.1, more than BA.4.6 and BA.2.75 combined. For all variants, those who experienced an outcome had a greater average age, with cases have an average age of 72.5 years, compared to controls with an average age of 56.8 years (difference 15.7, 95% CI: 14.8 to 16.7), and cases were more likely to be male than controls (51.4%, 95% CI: 50.0% to 52.7%). Across all variants, and for both cases and controls, more than half had had a vaccine greater than or equal to 3 months ago (7,018, 67.6%, Table 1).

**Table 1:** Demographic characteristics of cases and controls by Omicron sub-lineage.

|  | BA.4.6 (n = 432) |  | BA.2.75 (n = 527) |  | BQ.1 (n = 1,564) |  | BA.5 (n = 7,852) |  |
| --- | --- | --- | --- | --- | --- | --- | --- | --- |
|  | Cases (%)<br>(n = 208) | Controls (%)<br>(n = 224) | Cases (%) (n<br>= 252) | Controls (%) (n<br>= 275) | Cases (%) (n<br>= 784) | Controls (%) (n<br>= 780) | Cases (%) (n<br>= 3,963) | Controls (%) (n<br>= 3,889) |
| **Age group** |  |  |  |  |  |  |  |  |
| 0-9 | 4 (1.9) | 25 (11.2) | 5 (2.0) | 25 (9.1) | 25 (3.2) | 69 (8.8) | 120 (3.0) | 324 (8.3) |
| 10-19 | 2 (1.0) | 6 (2.7) | 3 (1.2) | 8 (2.9) | 10 (1.3) | 18 (2.3) | 23 (0.6) | 102 (2.6) |
| 20-29 | 4 (1.9) | 24 (10.7) | 7 (2.8) | 24 (8.7) | 13 (1.7) | 59 (7.6) | 63 (1.6) | 310 (8.0) |
| 30-39 | 10 (4.8) | 21 (9.4) | 6 (2.4) | 22 (8.0) | 18 (2.3) | 75 (9.6) | 88 (2.2) | 349 (9.0) |
| 40-49 | 6 (2.9) | 31 (13.8) | 6 (2.4) | 20 (7.3) | 19 (2.4) | 50 (6.4) | 129 (3.2) | 281 (7.2) |
| 50-59 | 10 (4.8) | 17 (7.6) | 17 (6.7) | 37 (13.4) | 52 (6.6) | 90 (11.5) | 281 (7.1) | 367 (9.4) |
| 60-69 | 27 (13.0) | 20 (8.9) | 30 (11.9) | 26 (9.5) | 93 (11.9) | 78 (10.0) | 475 (12.0) | 502 (12.9) |
| 70-79 | 47 (22.6) | 31 (13.8) | 61 (24.2) | 53 (19.3) | 180 (23.0) | 142 (18.2) | 1,025 (25.9) | 668 (17.2) |
| 80-89 | 67 (32.2) | 37 (16.5) | 92 (36.5) | 42 (15.3) | 278 (35.5) | 135 (17.3) | 1,209 (30.5) | 704 (18.1) |
| ≥ 90 | 31 (14.9) | 12 (5.4) | 25 (9.9) | 18 (6.5) | 96 (12.2) | 64 (8.2) | 550 (13.9) | 282 (7.3) |
| **Sex** |  |  |  |  |  |  |  |  |
| Female | 102 (49.0) | 134 (59.8) | 117 (46.4) | 136 (49.5) | 383 (48.9) | 432 (55.4) | 1,930 (48.7) | 2,008 (51.6) |
| Male | 106 (51.0) | 90 (40.2) | 135 (53.6) | 139 (50.5) | 401 (51.1) | 348 (44.6) | 2,033 (51.3) | 1,881 (48.4) |
| **Vaccination status** |  |  |  |  |  |  |  |  |
| 0-2 weeks | 9 (4.3) | 9 (4.0) | 16 (6.3) | 14 (5.1) | 65 (8.3) | 50 (6.4) | 225 (5.7) | 177 (4.6) |
| 3 weeks-<3 months | 28 (13.5) | 21 (9.4) | 46 (18.3) | 34 (12.4) | 191 (24.4) | 191 (24.5) | 427 (10.8) | 332 (8.5) |
| ≥ 3 months | 152 (73.1) | 145 (64.7) | 161 (63.9) | 172 (62.5) | 453 (57.8) | 413 (52.9) | 2,887 (72.8) | 2,635 (67.8) |
| Unvaccinated | 19 (9.1) | 49 (21.9) | 29 (11.5) | 55 (20.0) | 75 (9.6) | 126 (16.2) | 424 (10.7) | 745 (19.2) |
| **Index of multiple deprivation quintile** |  |  |  |  |  |  |  |  |
| 1 (most deprived) | 61 (29.3) | 58 (25.9) | 60 (23.8) | 72 (26.2) | 208 (26.5) | 177 (22.7) | 930 (23.5) | 912 (23.5) |
| 2 | 36 (17.3) | 53 (23.7) | 50 (19.8) | 68 (24.7) | 143 (18.2) | 163 (20.9) | 733 (18.5) | 871 (22.4) |
| 3 | 33 (15.9) | 53 (23.7) | 53 (21.0) | 53 (19.3) | 143 (18.2) | 157 (20.1) | 777 (19.6) | 793 (20.4) |
| 4 | 39 (18.8) | 30 (13.4) | 47 (18.7) | 45 (16.4) | 152 (19.4) | 149 (19.1) | 809 (20.4) | 699 (18.0) |
| 5 (least deprived) | 39 (18.8) | 30 (13.4) | 42 (16.7) | 37 (13.5) | 138 (17.6) | 134 (17.2) | 714 (18.0) | 614 (15.8) |
| **Region** |  |  |  |  |  |  |  |  |
| East of England | 9 (4.3) | 17 (7.6) | 24 (9.5) | 28 (10.2) | 69 (8.8) | 67 (8.6) | 383 (9.7) | 352 (9.1) |
| London | 15 (7.2) | 23 (10.3) | 38 (15.1) | 42 (15.3) | 93 (11.9) | 89 (11.4) | 309 (7.8) | 343 (8.8) |
| Midlands | 49 (23.6) | 41 (18.3) | 63 (25.0) | 70 (25.5) | 189 (24.1) | 204 (26.2) | 911 (23.0) | 1,040 (26.7) |
| North East and Yorkshire | 43 (20.7) | 25 (11.2) | 34 (13.5) | 27 (9.8) | 139 (17.7) | 79 (10.1) | 678 (17.1) | 440 (11.3) |
| North West | 48 (23.1) | 48 (21.4) | 36 (14.3) | 28 (10.2) | 109 (13.9) | 110 (14.1) | 606 (15.3) | 552 (14.2) |
| South East | 22 (10.6) | 41 (18.3) | 30 (11.9) | 50 (18.2) | 119 (15.2) | 143 (18.3) | 686 (17.3) | 704 (18.1) |
| South West | 22 (10.6) | 29 (12.9) | 27 (10.7) | 30 (10.9) | 66 (8.4) | 88 (11.3) | 390 (9.8) | 458 (11.8) |

A total of 6,417 episodes met the study criteria for Population 2 and were included in the analysis, of which 3,150 (49.1%) were admitted or died. Similar patterns were seen among variants and cases and controls as in the primary population (Supplemental Materials, Table 1, and Figure 1).

### Severity Results

There was no evidence for a difference in the odds of admission or death for BA.4.6 (OR=0.94, 95% CI: 0.82 to 1.08), BA.2.75 (OR=0.92, 95% CI: 0.80 to 1.05), and BQ.1 (OR= 1.02, 95% CI: 0.93 to 1.11) in an unadjusted analysis. Following adjustment for test week, age group, vaccination status and NHS region, there remained no evidence for greater odds of admission or death among those infected with BA.2.75 (OR 0.96, 95% CI: 0.84 to 1.09), BA.4.6 (OR 1.02, 95% CI: 0.88 to 1.17), or BQ.1 (OR= 1.03, 95% CI: 0.94 to 1.13) compared to BA.5. (Figure 2).

**Figure 2:**
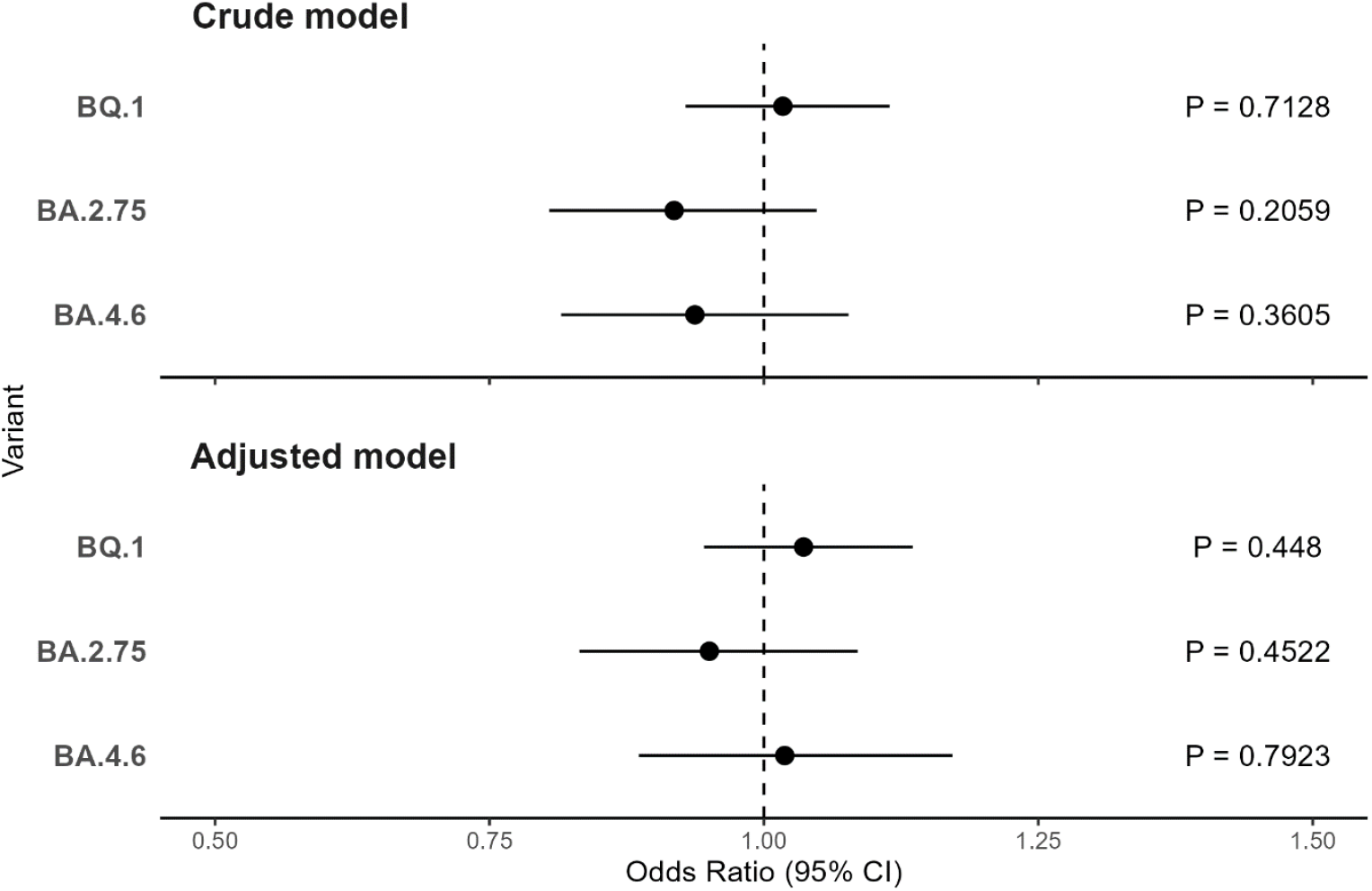
Crude and adjusted odds ratios for admission or death, relative to BA.5 with 95% Confidence Intervals of BA.4.6, BA.2.75 and BQ.1

The results for the Population 2, those who tested positive on the same day as attendance to the ED, were similar to those in the primary definition. Following adjustment, there was some evidence for greater odds of admission or death among those infected with BQ.1 (OR= 1.08, 95% CI: 0.95 to 1.22), however there was insufficient evidence to conclude that this was not a chance finding. Likewise, there remained no evidence for difference in the odds of admission or death among those infected with BA.2.75 (OR= 0.94, 95% CI: 0.79 to 1.11) and BA.4.6 (OR=1.02, 95% CI: 0.86 to 1.22) relative to BA.5 (Supplemental Materials Figure 2).

## Discussion

### Severity of BA.2.75, BA.4.6 and BQ.1 compared to BA.5

The results do not suggest a difference in risk of hospital admission or death, following attendance to emergency care, for individuals with BA.2.75, BA.4.6 or BQ.1 compared to individuals with BA.5. This follows similar findings seen among other Omicron sub-lineages of not showing evidence of being more severe than their predecessor. (19–21)

### Strengths and Limitations

This analysis benefitted from the large volumes of data available, including access to hospitalisation data in England and vaccination status. While previous estimates of relative severity of SARS-CoV-2 variants have used cohort studies in which all people testing positive for a given variant, the reduction in community testing overall in England has meant that a more targeted approach was needed. Whole genome sequencing was prioritised for high-risk groups such as care home residents, individuals eligible for COVID-19 therapeutics and people admitted to hospital. (22) For this reason, a case-control study design was used to assess the risk of admission for those presenting to the ED. With the change in testing implementation, there is a higher chance of hospital samples being sequenced, and the chosen population may reflect those who were already at a higher risk of hospital admission than the general population.

Presenting to emergency care is already a form of severe infection, therefore the controls include in the study are likely to be those whose infection is more severe than those infections occurring among the general population. This restriction of only including episodes among people presenting to emergency care may mean that the results obtained here are less generalisable to the general population. However, in the absence of an observed difference in risk between variants and a lack of a mechanism by which one variant might more severely affect the general population and not a high-risk population, any lack of generalisability is of low concern to the validity of these results.

The second population definition is likely more representative of the general population, as it includes individuals who tested positive on attendance to the ED, suggesting they have limited access to tests outside of healthcare or high-risk setting, indicating they are less likely to be pre-selected for anti-COVID-19 therapeutics. However, testing in this setting could be more sensitive to incidental COVID-19, in which COVID-19 is not the primary reason for attendance to emergency care. To investigate the sensitivity of our results to such factors we used both definitions in the analysis and observed comparable results between the two definitions.

With the nature of how the sub-lineages of Omicron evolved and emerged over time, there were a limited number of observations in which new variants occurred in temporal proximity with BA.5 episodes, somewhat limiting the power of this analysis. However, the analysis accounted for this by stratifying by week, which also captures difficult to measure confounding factors such as hospital capacity, health-seeking behaviours, and risk avoidance behaviour in at-risk populations. Although, the study was underpowered to detect the small difference in odds presented in the analysis, a further limitation of using surveillance data, the study was well powered to detect larger differences.

## Conclusion

The results described do not indicate that there is a difference in severity of illness between the previously dominant Omicron BA.5 sub-lineage and the subsequent Omicron BA.4.6 and BA.2.75 or BQ.1 sub-lineages. These findings will provide key insights to the public health management of Omicron variants in England.

However, future lineages may not follow the same trend seen with the analysis presented. There remains a need for continued surveillance of COVID-19 variants and sub-lineages and their clinical outcomes to inform the public health response to future emerging variants and sub-lineages.

## Acknowledgements

The authors acknowledge Tommy Nyberg, Anne Presanis and Daniela De Angelis from MRC Biostatistics Unit, University of Cambridge, for their expert advice.

## Author Contributions

ST and GD conceived and designed the study. GS prepared the datasets and performed the statistical analysis, supported by ST and NA. GS, ST, SN drafted the first version of the manuscript. All authors read, revised, and approved the final version of the manuscript.

## Financial Support

This research received no specific grant from any funding agency, commercial or not-for-profit sectors.

## Conflict of Interest

None.

## Data Availability Statement

The individual-level nature of the data used risks individuals being identified, or being able to self-identify, if the data are released publicly. Requests for access to these non-publicly available data should be directed to UKHSA.

## Ethics

UKHSA has legal permission, provided by Regulation 3 of The Health Service (Control of Patient Information) Regulations 2002 to process confidential patient information under Sections 3(i) (a) to (c), 3(i)(d) (i) and (ii) and 3(iii) as part of its outbreak response activities. This study falls within the research activities approved by the UKHSA Research Ethics and Governance Group.

## Notes

### Competing Interest Statement

The authors have declared no competing interest.

