## Supplemental Materials for "Risk of severe outcomes among Omicron sub-lineages BA.4.6, BA.2.75 and BQ.1 compared to BA.5 in England"

### Supplemental Material

Within UKHSA, SARS-CoV-2 lineages are usually assigned to sequences using Pangolin (Ultrafast Sample placement on Existing tRee (UShER) analysis engine). (1) This uses mutations present throughout the genome to place sequences in a phylogeny and assign a lineage. UKHSA define variants based on lineages of concern to allow consistent detection, monitoring and reporting. The definitions include a set of mutations relevant to the Wuhan-1 reference (NC_045512) that can be used in combination to distinguish the variant from other SARS-CoV-2 variants. A minimum number of mutations required to provide sufficient sensitivity and specificity is determined. The list of mutations and calling requirements can be found on the UKHSA variant definitions repository.(2) Any sequences that do not meet these criteria will not be reported in the variant counts regardless of whether there is other evidence to suggest that the sequence belongs to the relevant pangolin lineage. Sequences that do not meet the current UKHSA definition but are assigned by pangolin to a relevant lineage are reviewed to ensure the suitability of these variant definitions over time.

1. O'Toole Á, Scher E, Underwood A, Jackson B, Hill V, McCrone JT, et al. Assignment of epidemiological lineages in an emerging pandemic using the pangolin tool. Virus Evol. 2021 Dec 16;7(2):veab064.

2. Matt Bull, Meera Chand, Tom Connor, Nick Ellaby, Natalie Groves. Standardised Variant Definitions [Internet]. UKHSA-genomics; 2022 [cited 2022 Dec 23]. Available from: https://github.com/ukhsa-collaboration/variant_definitions

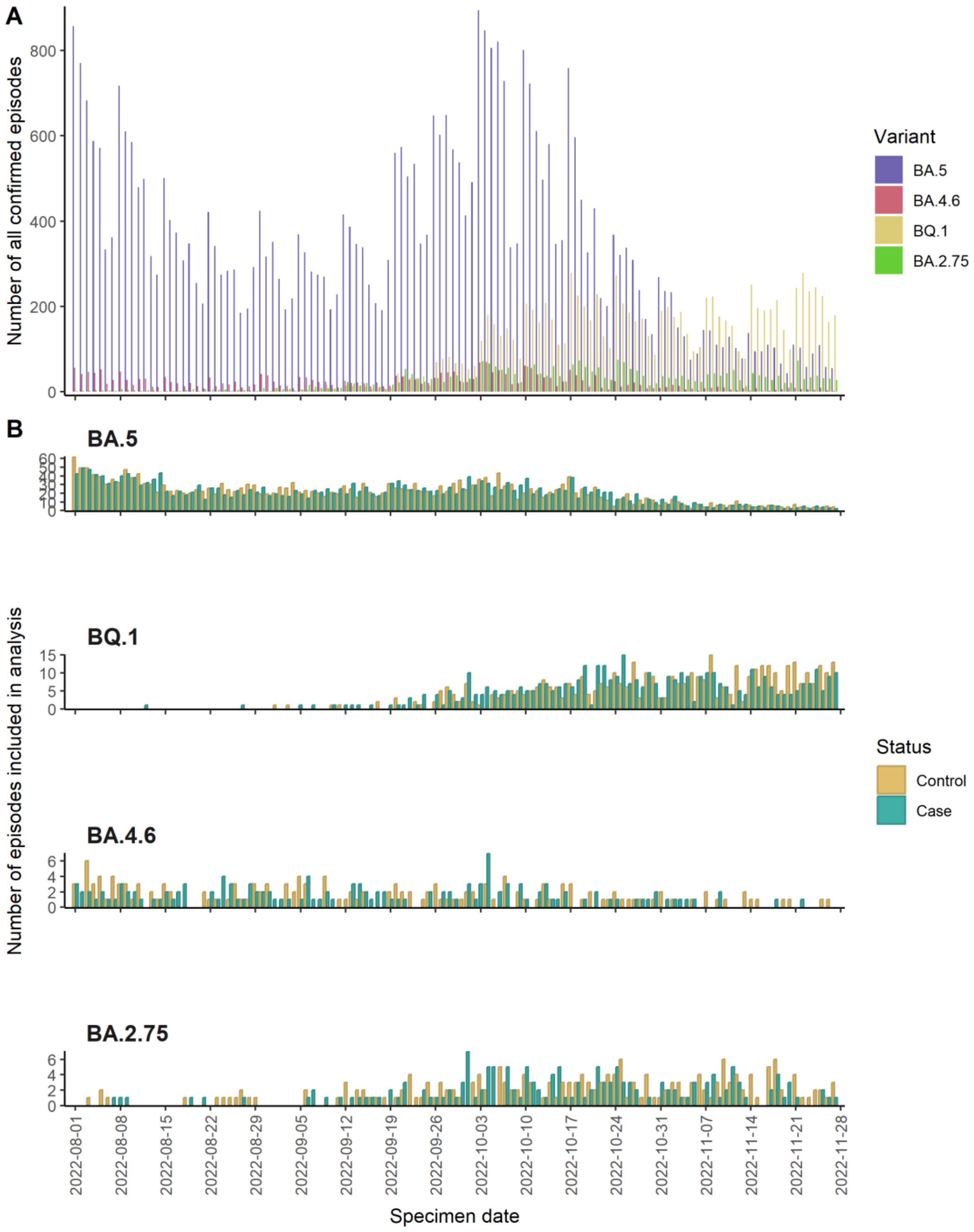


*Figure 1: Epicurves of cases and controls for BA.5, BA.4.6, BA.2.75 and BQ.1 for Population 2*

| | BA.4.6 (n = 288) | | BA.2.75 (n = 355) | | BQ.1 (n = 878) | | BA.5 (n = 4,896) | |
|---|---|---|---|---|---|---|---|---|
| | Cases (%) (n = 137) | Controls (%) (n = 151) | Cases (%) (n = 160) | Controls (%) (n = 195) | Cases (%) (n = 437) | Controls (%) (n = 441) | Cases (%) (n = 2,416) | Controls (%) (n = 2,480) |
| **Age group** | | | | | | | | |
| 0-9 | 2 (1.5) | 17 (11.3) | 3 (1.9) | 18 (9.2) | 11 (2.5) | 44 (10.0) | 80 (3.3) | 240 (9.7) |
| 10-19 | 2 (1.5) | 4 (2.6) | 2 (1.2) | 7 (3.6) | 8 (1.8) | 8 (1.8) | 16 (0.7) | 70 (2.9) |
| 20-29 | 1 (0.7) | 19 (12.6) | 4 (2.5) | 17 (8.7) | 6 (1.4) | 36 (8.2) | 44 (1.8) | 215 (8.7) |
| 30-39 | 9 (6.6) | 13 (8.6) | 4 (2.5) | 18 (9.2) | 13 (3.0) | 52 (11.8) | 49 (2.0) | 248 (10.0) |
| 40-49 | 4 (2.9) | 24 (15.9) | 3 (1.9) | 16 (8.2) | 13 (3.0) | 25 (5.7) | 76 (3.1) | 172 (6.9) |
| 50-59 | 6 (4.4) | 11 (7.3) | 11 (6.9) | 28 (14.4) | 37 (8.5) | 52 (11.8) | 171 (7.1) | 242 (9.8) |
| 60-69 | 14 (10.2) | 14 (9.3) | 23 (14.4) | 22 (11.3) | 54 (12.4) | 42 (9.5) | 280 (11.6) | 338 (13.6) |
| 70-79 | 40 (29.2) | 17 (11.3) | 35 (21.9) | 34 (17.4) | 94 (21.5) | 82 (18.6) | 633 (26.2) | 407 (16.4) |
| 80-89 | 41 (29.9) | 26 (17.2) | 55 (34.4) | 26 (13.3) | 155 (35.5) | 68 (15.4) | 741 (30.7) | 401 (16.2) |
| ≥ 90 | 18 (13.1) | 6 (4.0) | 20 (12.5) | 9 (4.6) | 46 (10.5) | 32 (7.3) | 326 (13.5) | 147 (5.9) |
| **Sex** | | | | | | | | |
| Female | 64 (46.7) | 94 (62.3) | 73 (45.6) | 98 (50.3) | 207 (47.4) | 240 (54.4) | 1,176 (48.7) | 1,263 (50.9) |
| Male | 73 (53.3) | 57 (37.7) | 87 (54.4) | 97 (49.7) | 230 (52.6) | 201 (45.6) | 1,240 (51.3) | 1,217 (49.1) |
| **Vaccination status** | | | | | | | | |
| 0-2 weeks | 8 (5.8) | 5 (3.3) | 10 (6.2) | 9 (4.6) | 37 (8.5) | 26 (5.9) | 141 (5.8) | 101 (4.1) |
| 3 weeks-<3 months | 17 (12.4) | 11 (7.3) | 29 (18.1) | 21 (10.8) | 103 (23.6) | 98 (22.2) | 234 (9.7) | 168 (6.8) |
| >= 3 months | 96 (70.1) | 100 (66.2) | 99 (61.9) | 123 (63.1) | 258 (59.0) | 240 (54.4) | 1,755 (72.6) | 1,670 (67.3) |
| Unvaccinated | 16 (11.7) | 35 (23.2) | 22 (13.8) | 42 (21.5) | 39 (8.9) | 77 (17.5) | 286 (11.8) | 541 (21.8) |
| **Index of multiple deprivation quintile** | | | | | | | | |
| 1 | 41 (29.9) | 35 (23.2) | 36 (22.5) | 51 (26.2) | 105 (24.0) | 92 (20.9) | 548 (22.7) | 579 (23.3) |
| 2 | 21 (15.3) | 36 (23.8) | 30 (18.8) | 49 (25.1) | 87 (19.9) | 100 (22.7) | 461 (19.1) | 574 (23.1) |
| 3 | 24 (17.5) | 40 (26.5) | 36 (22.5) | 39 (20.0) | 82 (18.8) | 95 (21.5) | 476 (19.7) | 498 (20.1) |
| 4 | 25 (18.2) | 18 (11.9) | 30 (18.8) | 31 (15.9) | 81 (18.5) | 81 (18.4) | 497 (20.6) | 438 (17.7) |
| 5 | 26 (19.0) | 22 (14.6) | 28 (17.5) | 25 (12.8) | 82 (18.8) | 73 (16.6) | 434 (18.0) | 391 (15.8) |
| **Region** | | | | | | | | |
| East of England | 5 (3.6) | 13 (8.6) | 18 (11.2) | 21 (10.8) | 46 (10.5) | 40 (9.1) | 231 (9.6) | 225 (9.1) |
| London | 12 (8.8) | 18 (11.9) | 29 (18.1) | 33 (16.9) | 60 (13.7) | 53 (12.0) | 213 (8.8) | 235 (9.5) |
| Midlands | 31 (22.6) | 29 (19.2) | 35 (21.9) | 57 (29.2) | 99 (22.7) | 117 (26.5) | 539 (22.3) | 696 (28.1) |
| North East and Yorkshire | 26 (19.0) | 12 (7.9) | 14 (8.8) | 15 (7.7) | 65 (14.9) | 31 (7.0) | 333 (13.8) | 230 (9.3) |
| North West | 30 (21.9) | 29 (19.2) | 23 (14.4) | 21 (10.8) | 58 (13.3) | 53 (12.0) | 362 (15.0) | 333 (13.4) |
| South East | 17 (12.4) | 28 (18.5) | 25 (15.6) | 27 (13.8) | 66 (15.1) | 84 (19.0) | 471 (19.5) | 461 (18.6) |
| South West | 16 (11.7) | 22 (14.6) | 16 (10.0) | 21 (10.8) | 43 (9.8) | 63 (14.3) | 267 (11.1) | 300 (12.1) |

*Table 1: Demographic characteristics of cases and controls by Omicron sub-lineage for Population 2*

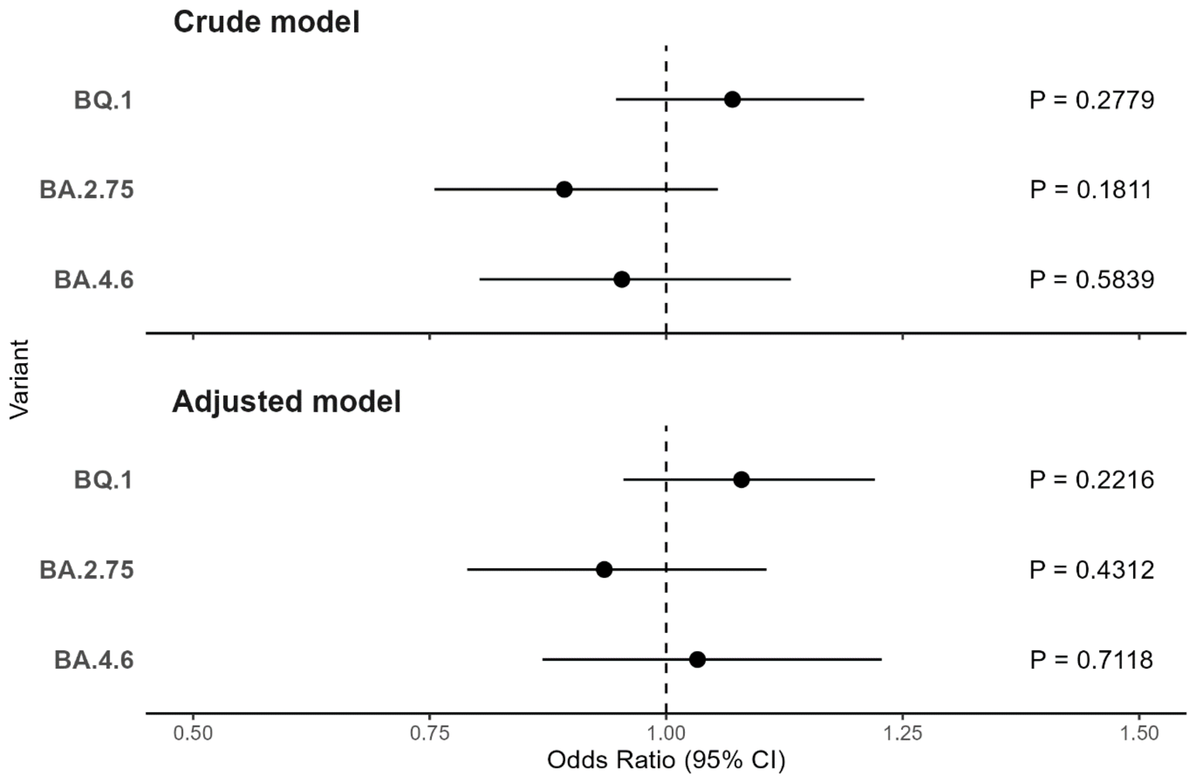


*Figure 2: Crude and Adjusted Odds Ratios with 95% Confidence Intervals of BA.4.6, BA.2.75 and BQ.1 for Population 2*
